# Detection and genome sequencing of SARS-CoV-2 belonging to the B.1.1.7 lineage in the Philippines

**DOI:** 10.1101/2021.03.04.21252557

**Authors:** Francis A. Tablizo, Cynthia P. Saloma, Marc Jerrone R. Castro, Kenneth M. Kim, Maria Sofia L. Yangzon, Carlo M. Lapid, Benedict A. Maralit, Marc Edsel C. Ayes, Jan Michael C. Yap, Jo-Hannah S. Llames, Shiela Mae M. Araiza, Kris P. Punayan, Irish Coleen A. Asin, Candice Francheska B. Tambaoan, Asia Louisa U. Chong, Karol Sophia Agape R. Padilla, Rianna Patricia S. Cruz, El King D. Morado, Joshua Gregor A. Dizon, Eva Maria Cutiongco-de la Paz, Alethea R. de Guzman, Razel Nikka M. Hao, Arianne A. Zamora, Devon Ray Pacial, Juan Antonio R. Magalang, Marissa Alejandria, Celia Carlos, Anna Ong-Lim, Edsel Maurice Salvaña, John Q. Wong, Jaime C. Montoya, Maria Rosario Singh-Vergeire

## Abstract

We report the sequencing and detection of 39 Severe Acute Respiratory Syndrome Coronavirus 2 (SARS-CoV-2) samples containing lineage-defining mutations specific to viruses belonging to the B.1.1.7 lineage (UK variant) in the Philippines.

## ANNOUNCEMENT

The Coronavirus Disease 2019 (COVID-19) is an infectious disease that has gained pandemic status from the World Health Organization, with millions of cases and deaths recorded worldwide. This global health crisis is caused by viruses referred to as Severe Acute Respiratory Syndrome Coronavirus 2 (SARS-CoV-2), a member of the genus *Betacoronavirus* (Coronaviridae) together with the causative agents of the first SARS outbreak in 2003 and the Middle East Respiratory Syndrome (MERS) in 2012.

In this study, we present the genome sequences of 39 cases of COVID-19 in the Philippines caused by viruses belonging to SARS-CoV-2 lineage B.1.1.7, more commonly known as the United Kingdom (UK) variant. This particular SARS-CoV-2 variant was initially identified in the United Kingdom and has been reported to cause a surge of COVID-19 infections in the said country (Kirby, 2021). Initial studies also suggest that the B.1.1.7 viruses appear to have replicative advantage (Grabowski et al., 2021) and are more transmissible (Leung et al., 2021).

A total of 39 Philippine SARS-CoV-2 samples were classified under the B.1.1.7 lineage. Table 1 shows the primary consensus assembly metrics for these samples. The average depth of coverage across all the sequences is 1,128x, with 33 of the samples carrying all 17 hallmark mutations of the B.1.1.7 lineage as listed in the PANGO Lineages report for the B.1.1.7 variant of concern (https://cov-lineages.org/global_report_B.1.1.7.html).

**Table 1.**
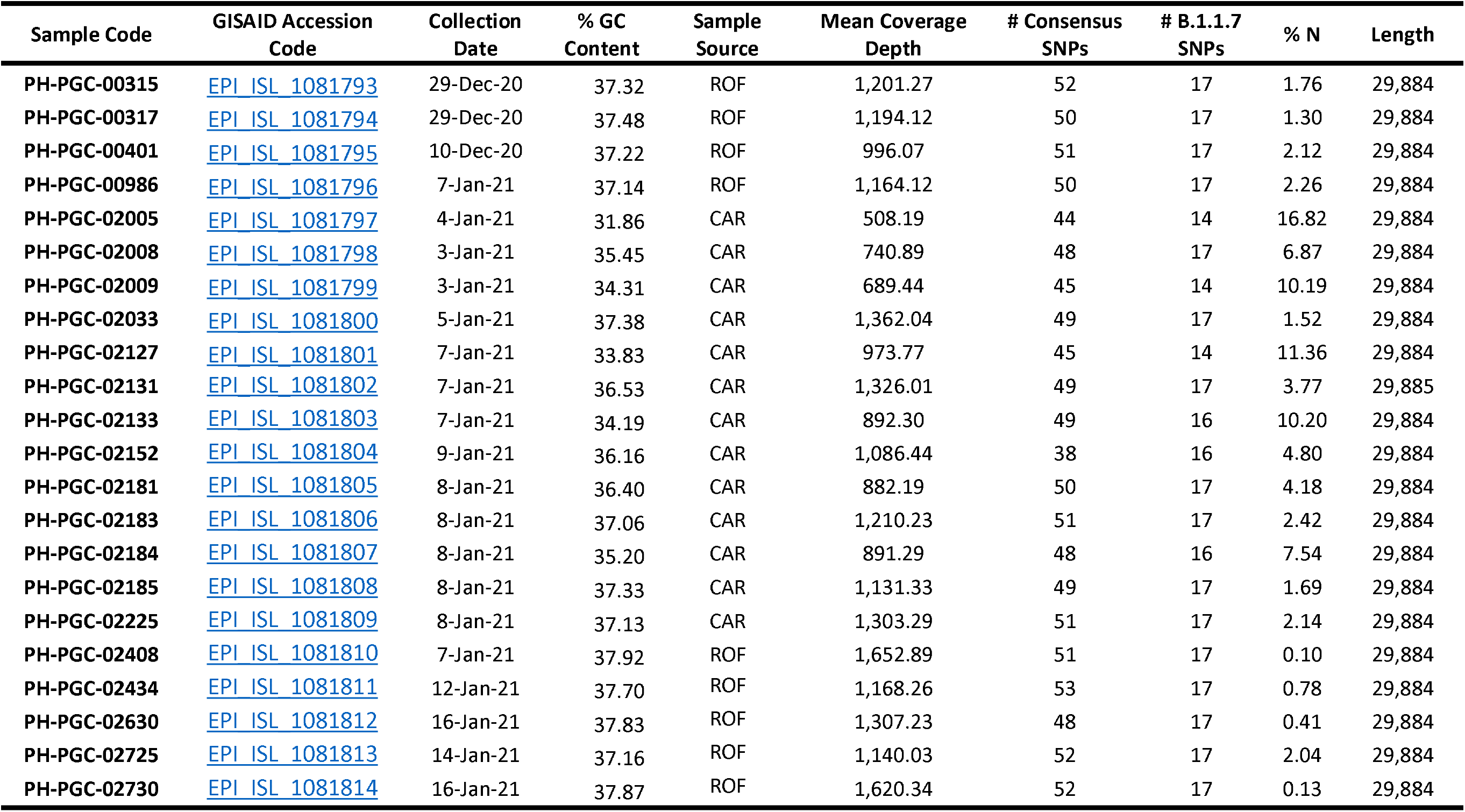
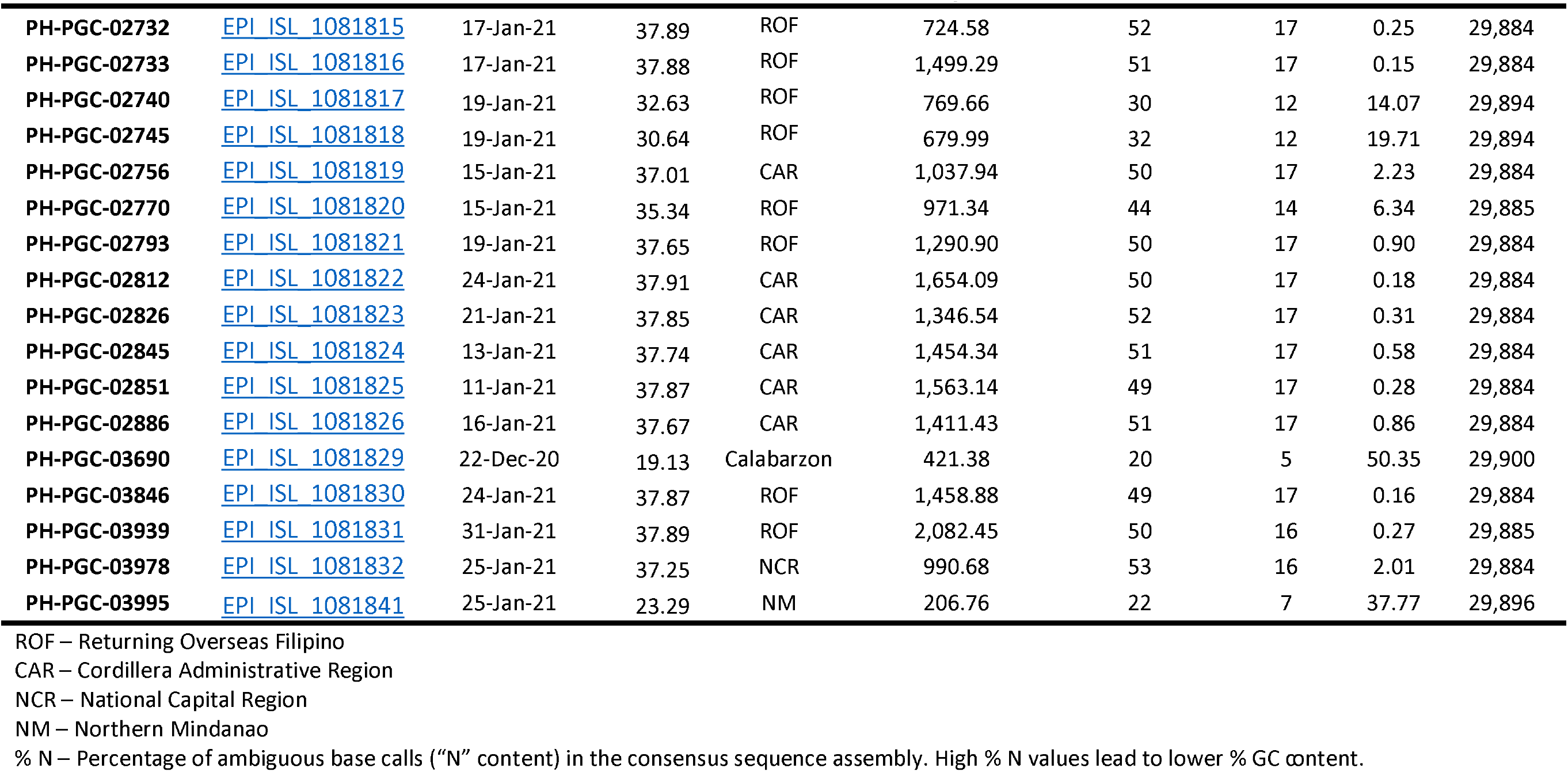
Primary consensus sequence assembly metrics.

The detection of B.1.1.7 from returning overseas Filipino workers and in the community highlights the need for genomic surveillance at the country’s ports of entry and the general population to monitor the importation and local transmission of emerging variants of concern that may impact the public health response to the SARS-CoV-2 pandemic in the Philippines.

## METHODS

In order to detect the entry of B.1.1.7 infection in the Philippines, nasopharyngeal swabs were collected between December 10, 2020 and January 31, 2021 from COVID-19 cases detected in returning overseas Filipinos, as well as from local case clusters mainly from the Cordillera Administrative Region of the country, among others. The collected swab samples were subjected to RNA extraction using the QIAamp Viral RNA Mini Kit, the product of which were then used as templates for the amplicon-based Illumina COVIDSeq Test (https://www.illumina.com/products/by-type/ivd-products/covidseq.html) sequencing workflow. Afterwards, the resulting sequence reads were mapped to the reference SARS-CoV-2 genome (NCBI Accession Number <u>NC 045512.2</u>) using Minimap2 (Li, 2018) version 2.17-r941, with the `–x sr` parameter for short accurate genomic reads alignment.

Primer clipping and quality trimming, intrahost variant calling, removal of reads associated with mismatched primer indices, and consensus sequence assembly were all done following the suggested workflow of iVar (Grubaugh et al., 2019) version 1.2.2, using default parameters. The consensus variants were identified by comparing the resulting assemblies with the reference sequence using MUMmer (Kurtz et al., 2004) as implemented in the RATT software (Otto et al., 2011). Lastly, SARS-CoV-2 lineage classifications were assigned using Pangolin version 2.3.2 (<u>github.com/cov-lineages/pangolin</u>; Rambaut et al., 2020).

## PUBLIC AVAILABILITY AND ACCESSION NUMBERS

The consensus sequence assemblies reported in this study are deposited in the <u>EpiCoV™ database of the GISAID</u>. The accession codes are shown in **Table 1**.

## Data Availability

The consensus sequence assemblies reported in this study are deposited in the EpiCoV database of the GISAID. The accession codes are shown in Table 1.

## ACKNOWLEDGEMENT

This project was supported by a Genomics Biosurveillance grant from the Philippine Department of Health, and a Department of Science and Technology – Philippine Council for Health Research and Development grant to BM and the University of the Philippines. The authors would also like to thank the contributing institutions to the Philippine Genomic Biosurveillance Network.

The protocols used in this study were reviewed and approved by the Single Joint Research Ethics Board of the Department of Health, with approval code SJREB-2021-11, as part of a larger research program entitled “A retrospective study on the national genomic surveillance of COVID-19 transmission in the Philippines by SARS-CoV-2 genome sequencing and bioinformatics analysis”.

